# A study of the visualization of artificial intelligence applications in chronic kidney disease in the literature over the last 20 years

**DOI:** 10.1101/2024.07.10.24310252

**Authors:** Yudi Li, Ying Ding, Yan Xu, Haoji Meng, Hongji Wu, Donglin Li, Yibo Hu

## Abstract

Chronic kidney disease (CKD) is a global public health problem characterized by persistent kidney damage or loss of kidney function. Previously, the diagnosis of CKD has mainly relied on serum creatinine and estimation of the glomerular filtration rate. However, with the development and progress of artificial intelligence (AI), AI has played different roles in various fields, such as early diagnosis, progression prediction, prediction of associated risk factors, and drug safety and efficacy evaluation. Therefore, research related to the application of AI in the field of CKD has become a hot topic at present. Therefore, this study adopts a bibliometric approach to study and analyze the development and evolution patterns and research hotspots of AI-CKD. English publications related to the field between January 1, 2004, and June 27, 2024, were extracted from the Web of Science Core Collection database. The research hotspots and trends of AI-CKD were analyzed at multiple levels, including publication trends, authors, institutions, countries, references and keywords, using VOSviewer and CiteSpace. The results showed that a total of 203 publications on AI-CKD were included in the study, of which Barbieri Carlo from the University of Milan, Italy, had the highest number of publications (NP=5) and had a high academic impact (H-Index=5), while the USA and its institution, the Mayo Clinic, were the publications. The USA and its Mayo Clinic are the countries and institutions with the highest number of publications, and China is the country with the second highest number of publications, with three institutions attributed to China among the top five institutions. Germany’s institution, Fresenius Medical Care, has the highest academic impact (H-index=6). Keyword analysis yielded artificial intelligence, chronic kidney disease, machine learning, prediction model, risk, deep learning, and other keywords with high frequency, and cluster analysis based on the timeline yielded a total of 8 machine learning, deep learning, retinal microvascular abnormality, renal failure, Bayesian network, anemia, bone disease, and allograft nephropathology clusters. This study provides a comprehensive overview of the current state of research and global frontiers of AI-CKD through bibliometric analysis. These findings can provide a valuable reference and guidance for researchers.

## 1. Introduction

Chronic kidney disease (CKD) is a global public health problem characterized by persistent kidney damage or loss of renal function. The diagnosis of CKD relies heavily on the assessment of renal function, including the measurement of serum creatinine (SCr) and the estimation of the glomerular filtration rate (eGFR) ^[1]^. The CKD-Epidemiology Collaboration (CKD-EPI) equation, which is based on the artificial intelligence (AI) algorithm, can more accurately estimate the GFR than can the traditional Modification of Diet in Renal Disease (MDRD) equation and can predict the risk of end-stage renal disease (ESRD) ^[1]^. With the development of AI in recent years, the application of AI has expanded from simple data processing to complex clinical decision support systems, providing more accurate and personalized healthcare services for CKD patients. However, there are no bibliometric studies analyzing AI in the field of CKD, and the current status of the field and the current research hotspots are not yet clear. To provide a reference for further research and application, this study used VOSviewer 1.6.20 and CiteSpace 6.3. R1 (Advanced) software was used to visualize and analyze the number of annual publications, authors, institutions, countries, journals, cited literature, keywords, and other aspects related to this field in the past two decades and to illustrate the research hotspots and cutting-edge trends of AI-CKD. research hotspots and cutting-edge development trends for future research.

## 2. Materials and Methods

### 2.1. Search Strategies

The data for this study were obtained from the Web of Science Core Collection (WOSCC), which is one of the most comprehensive and authoritative literature databases available and is widely used for bibliometric analysis. The search was conducted on June 27, 2024, to collect and retrieve data from the Science Citation Index Expanded (SCI-Expanded) and Social Sciences Citation Index (SSCI) citation index databases with a search formula. The search formula was as follows: (TS=(Artificial Intelligence)) AND TS=(Chronic kidney disease), and the Index Date was set from January 1, 2004, to June 27, 2024. Only Articles and Review Articles with English as the main language were included in this study, and Proceeding Paper, Book Chapters and Retracted Publication were excluded. A total of 203 papers were ultimately included, and the screening process is shown in Table 1.

**Table 1.** Relevant Literature Screening Process.

| Rank | Inclusion | Inclusion in the exclusion process |
| --- | --- | --- |
| 1 | 230 | Query: (TS=(Artificial Intelligence)) AND TS=(Chronic kidney disease) |
| 2 | 230 | Set year: Timespan: 2004-01-01 to 2024-06-27 (Index Date) |
| 3 | 210 | Set and select article type: Article or Review Article |
| 4 | 205 | Set and exclude article type: Proceeding Paper or Book Chapters or Retracted Publication |
| 4 | 203 | Set language: English |

### 2.2. Data Analysis

The data presented in this paper were processed and visualized using VOSviewer 1.6.20 and CiteSpace 6.3. R1 (Advanced) bibliometric software and Excel statistical software were used. The 203 documents were exported in the “Plain text file” format, and the content of the document records was “Full Record and Cited References.” The aforementioned documents were imported into VOSviewer and CiteSpace software. The annual trend statistics of publications were calculated with Excel statistical software. Based on VOSviewer, a bibliometric analysis of coauthors, institutions, countries, journals, and keywords was carried out. Based on CiteSpace, an emergence analysis, cluster analysis, and timeline graph visualization of the cited documents and keywords were conducted.

## 3. Results

### 3.1. Annual publication trends

A total of 203 publications related to the application of AI in CKD were retrieved from the Science Citation Index Expanded (WOSCC) database without duplicates. The number of publications (NP) from 2004 to 2024 is shown in Fig. 1. Figure 1 illustrates this trend. In 2005, an inaugural article on the subject was published, and from 2005 to 2018, related research was in its nascent stages, with a relatively low number of publications. The period from 2018 to 2019 was designated the outbreak phase (NP=14), while the subsequent period from 2020 to 2023 was designated the second outbreak phase, during which the highest number of publications was observed in 2023 (NP=66). Furthermore, based on the expected trend line of the annual issuance of cumulative articles derived from the correlation linear calculation, the cumulative number of publications from 2004 to 2024 shows a gradual upward trend (R^2^=0.9779), and the trend line follows a sixth-order polynomial distribution (Figure 1).

**Figure 1.**
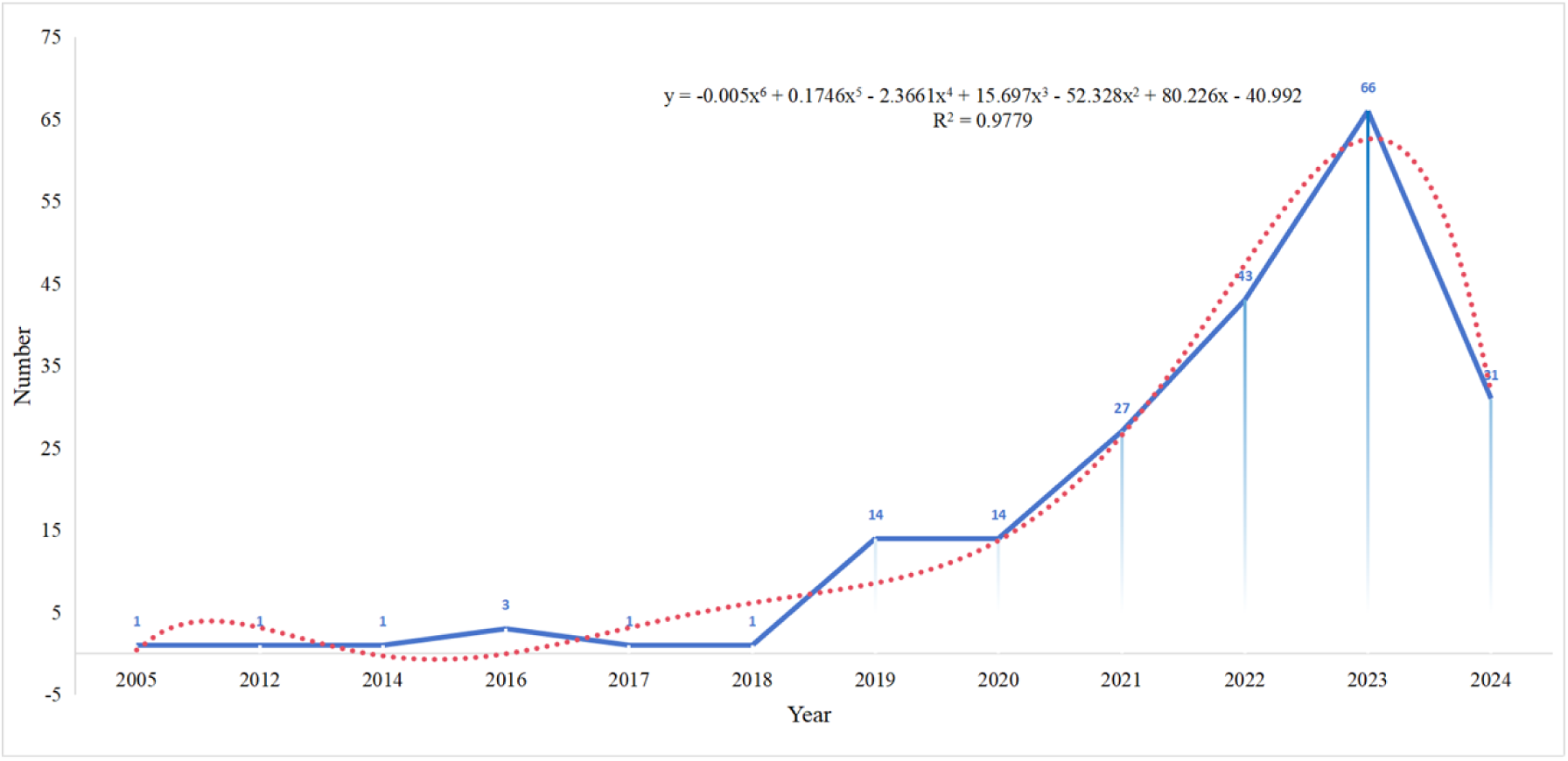
Annual number of publications and trends, 2004-2024.

### 3.2. Authors

A comparative analysis of the datasets in CiteSpace yielded 203 publications containing a total of 1,346 authors. VOSviewer was used to analyze and visualize the 46 authors with more than 100 publications cited (NCs) (Figure 2). The size of the circle indicates the number of publications, while the color gradient represents the average number of citations. The data pertaining to the top five authors of AI-related publications in the field of CKD were obtained. Among these, the research team led by Carlo Barbieri at the University of Milan, Italy, has a high academic impact, as evidenced by a high number of publications, citations, and H-indexes (Table **2**)

**Figure 2.**
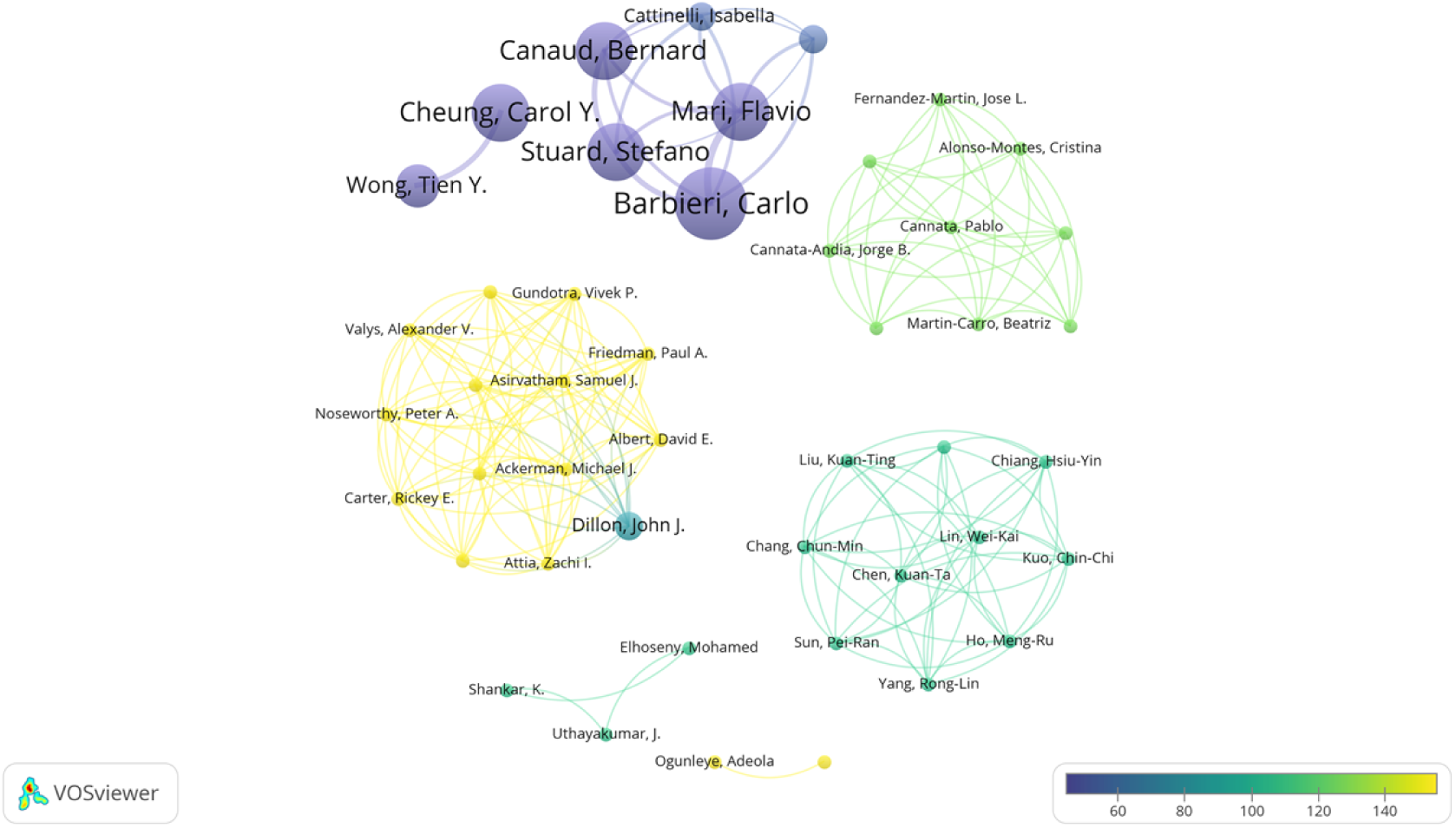
Author collaboration network.

**Table 2.** Institution NP and Academic Impact.

| Rank | Author | Institution | Country | NP | NC | H-index |
| --- | --- | --- | --- | --- | --- | --- |
| 1 | Barbieri Carlo | University of Milan | Italy | 5 | 195 | 5 |
| 2 | Canaud Bernard | University of Montpellier | Franch | 4 | 126 | 3 |
| 3 | Cheung, Carol Y. | The Chinese University of Hong Kong | China | 4 | 122 | 3 |
| 4 | Mari Flavio | Fresenius Med Care | Germany | 4 | 185 | 4 |
| 5 | Stuard Stefano | Fresenius Med Care | Germany | 4 | 136 | 4 |

### 3.3. Country and Institution

A total of 1,009 institutions and 52 countries were identified in the 203 documents. Of these, 21 countries (NP>5) and 39 institutions (NC>50) were subjected to analysis and visualization using VOSviewer (Figure 3). Figure 3a and Figure 3b illustrate the visualization results for countries and institutions, respectively. The analysis revealed that the number of publications and academic impact of AI in the field of CKD were greater in the USA and China (NP>50, H-index>10), followed by India and the UK (Table **3**). In terms of publications, the Mayo Clinic in Italy had the highest number of publications, while Fresenius Medical Care in Germany had a high number of citations and an H-index, indicating a significant academic impact in the field. Notably, three of the top five institutions are located in China (Table 4).

**Figure 3.**
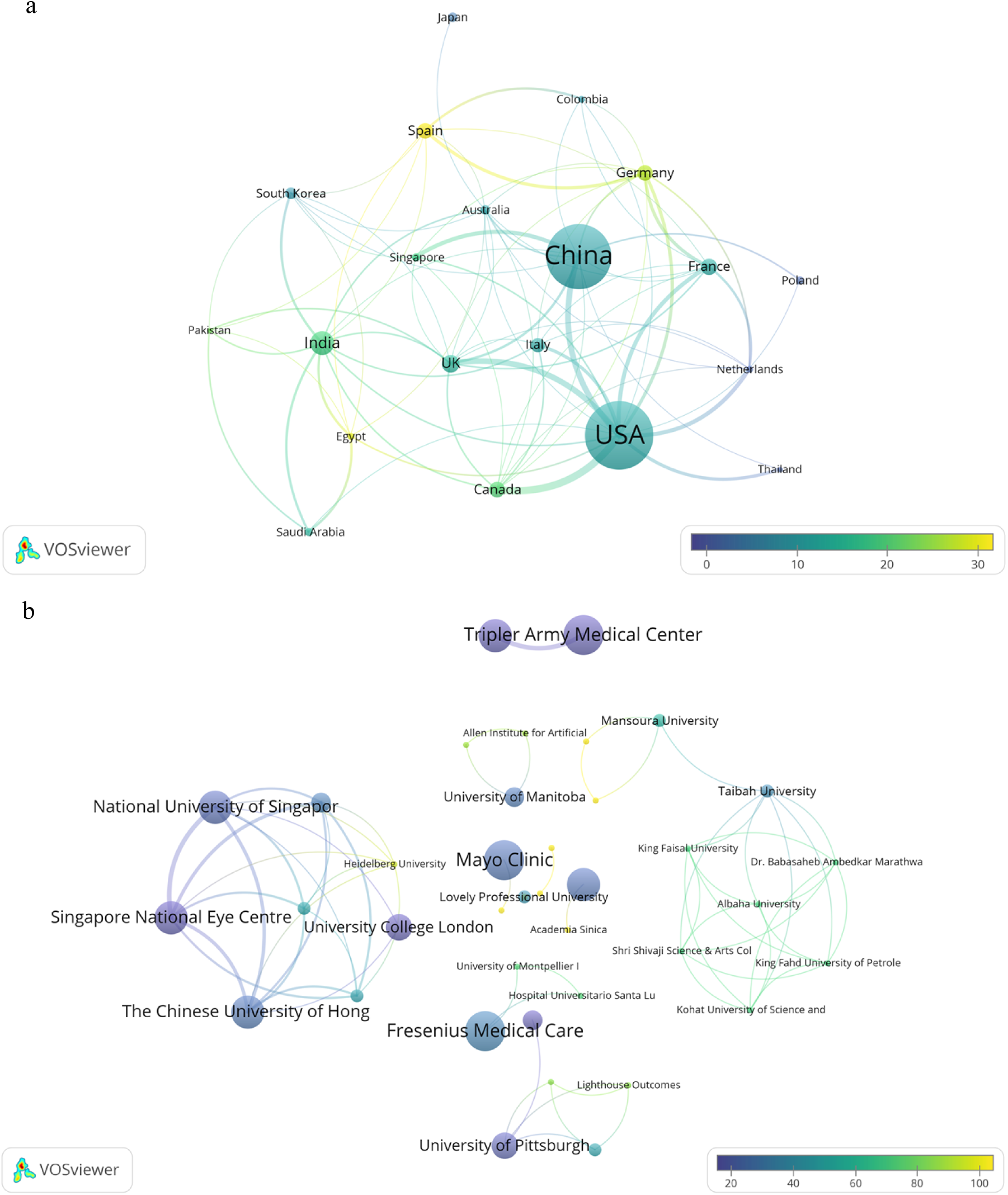
National and institutional cooperation networks.

**Table 3.** National NP and Academic Impact.

| Rank | Country | NP | NC | H-index | Average per item |
| --- | --- | --- | --- | --- | --- |
| 1 | USA | 57 | 647 | 12 | 11.35 |
| 2 | China | 54 | 564 | 12 | 10.44 |
| 3 | India | 20 | 370 | 9 | 18.5 |
| 4 | UK | 15 | 208 | 9 | 13.87 |
| 5 | France | 14 | 169 | 6 | 12.07 |

**Table 4.** Institutional NP and Academic Impact.

| Rank | Institution | Country | NP | NC | H-index | Average per item |
| --- | --- | --- | --- | --- | --- | --- |
| 1 | Mayo Clinic | USA | 7 | 170 | 3 | 24.29 |
| 2 | Fresenius Medical Care | Germany | 6 | 202 | 6 | 33.67 |
| 3 | Tri-Service General Hospital | China | 5 | 50 | 4 | 10.4 |
| 4 | China Medical University | China | 5 | 138 | 4 | 27.6 |
| 5 | The Chinese University of Hong Kong | China | 5 | 143 | 4 | 28.6 |

### 3.4. Keys

CiteSpace analysis yielded 203 documents containing 1,180 keywords. VOSviewer was used to analyze the co-occurrence of 42 keywords that appeared at least five times. This analysis revealed that the keywords “Artificial Intelligence,” “Chronic Kidney Disease,” “Machine Learning,” “Prediction Model,” “Risk,” “Deep Learning,” and other keywords are particularly prevalent (**Figure 5**b). A timeline-based cluster analysis was performed using CiteSpace, which resulted in the identification of eight sets of clusters, from #0 to #7, in the following order: machine learning, deep learning, retinal microvascular abnormality, renal failure, Bayesian network, anemia, bone diseases, and allograft nephropathology (**Figure 4**).

**Figure 4.**
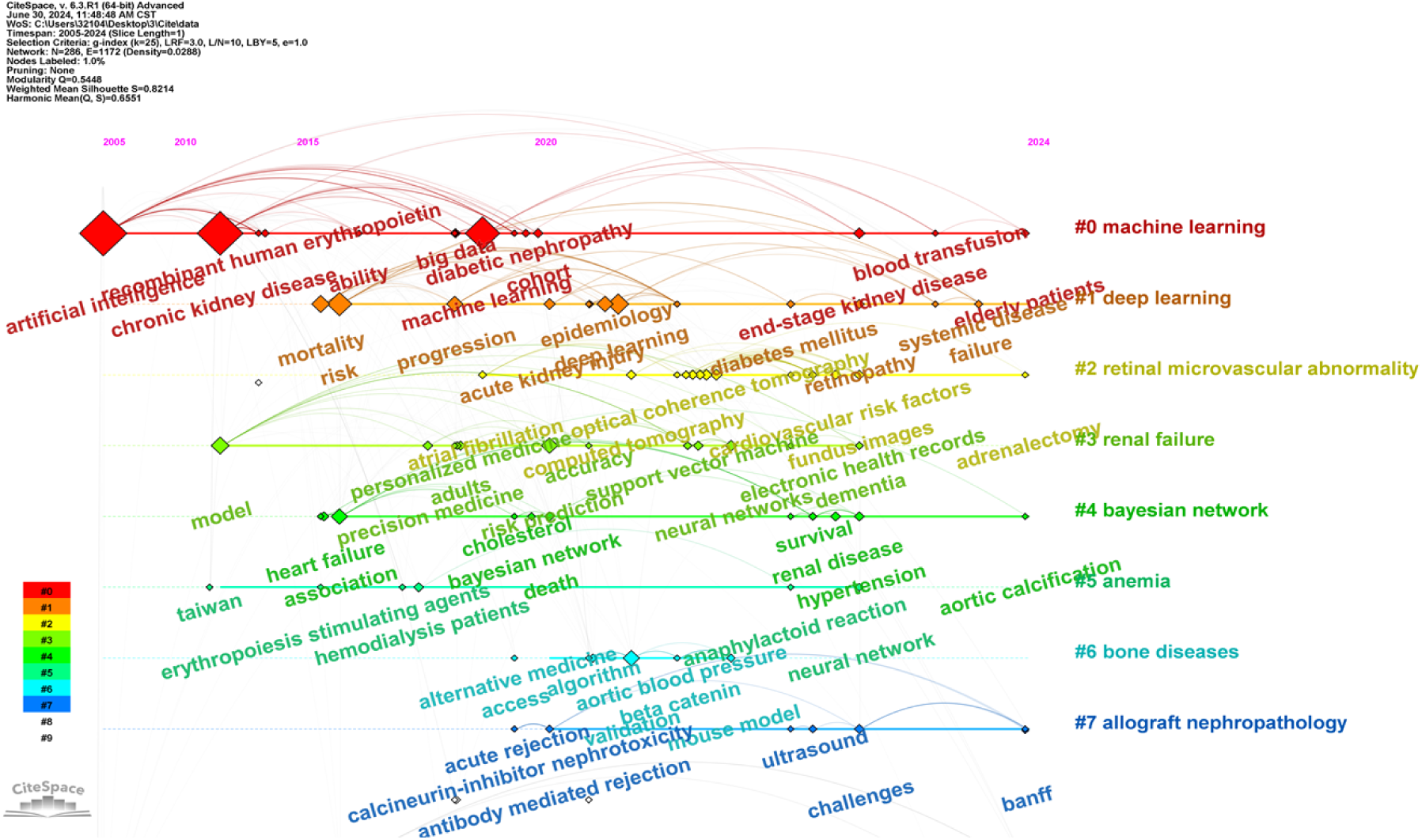
Time line graph co-occurrence analysis.

**Figure 5.**
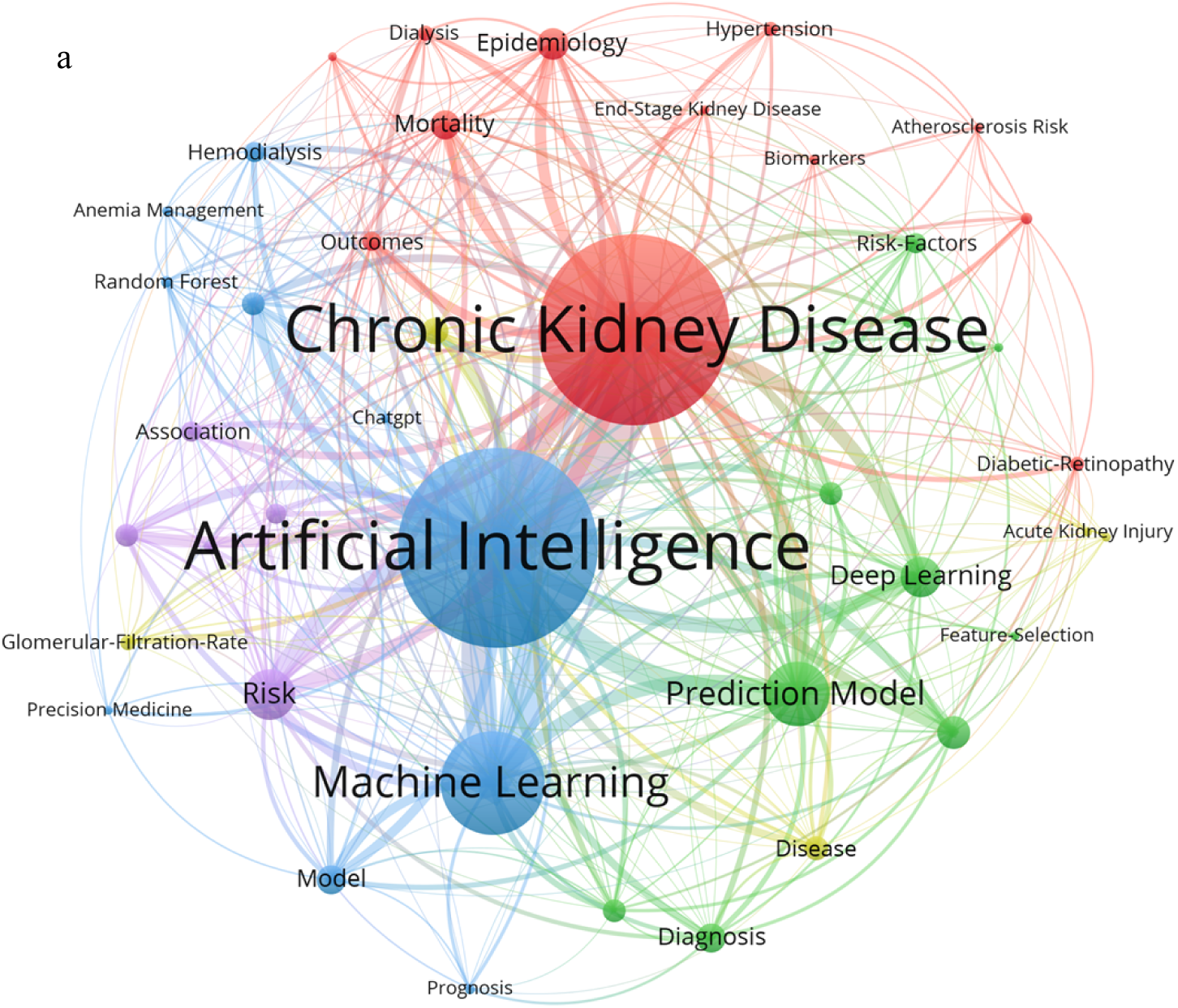
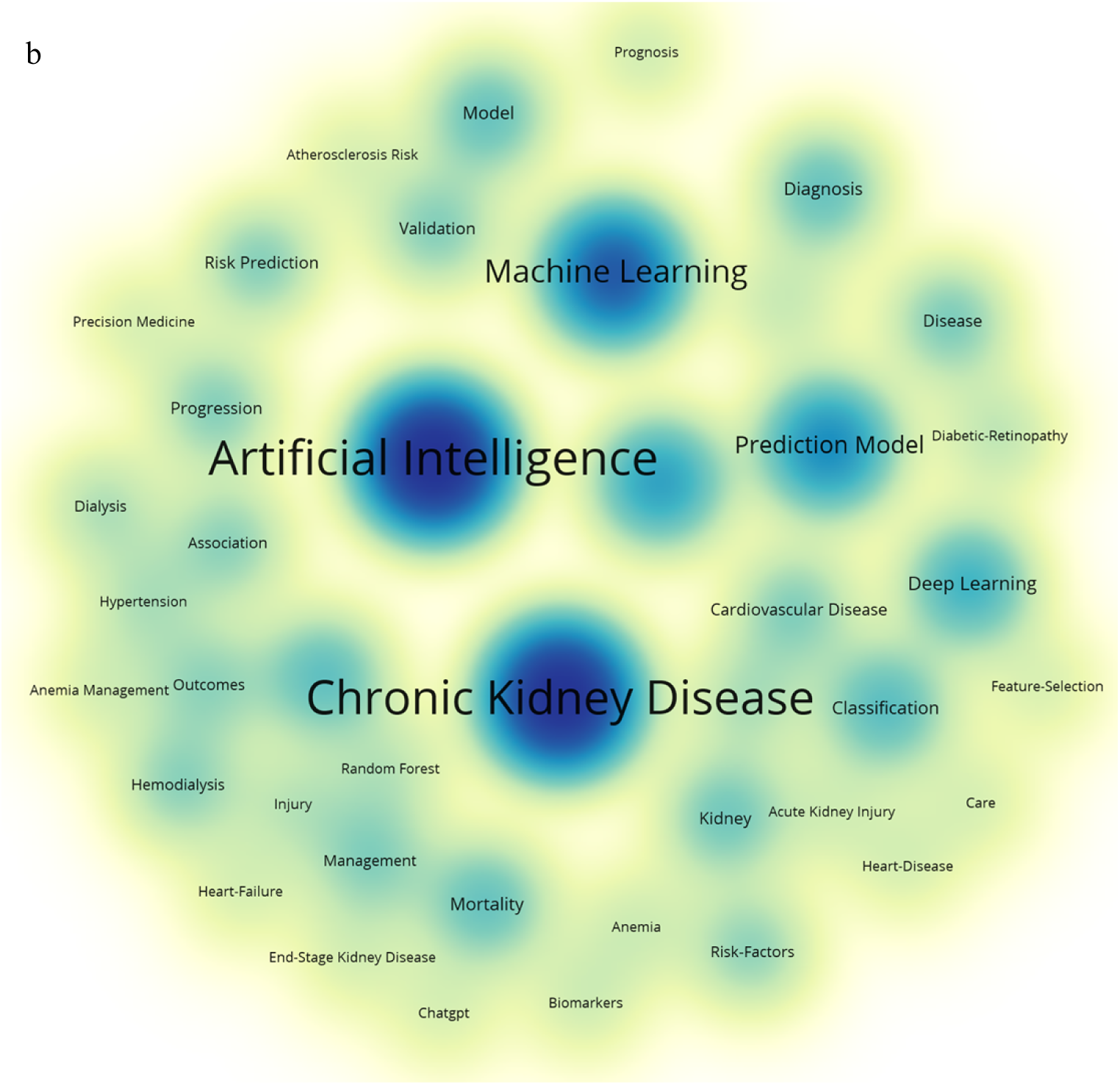
Keyword co-occurrence and density analysis.

## 4. Discussion

In 2005, the first study on the application of AI in the field of CKD was published. In this study, Marshall MR’s team developed a novel computational architecture based on AI, evolving connectionist systems (ECOS), to predict GFR in CKD patients, which is more accurate and smarter than the previous use of the MDRD formula to predict GFR. The results of the study show that ECOS has a deviation value of -0.1 in predicting the GFR after optimization, while the MDRD formula has a deviation value of -1.2 after optimization ^[2]^. During the period of 2005-2018, there were fewer related studies in this field, and it was still in its infancy. The period from 2018–2019 was the first research explosion phase, with a total of 14 publications for the whole year of 2019. The period from 2020-2023 was the second research explosion phase, and the number of publications increased annually, with a total of 66 publications in 2023, which was the year with the highest number of publications. As of June 27, 2024, the number of annual publications in this field of research reached 31, with a cumulative total of 203 (Figure 1).

The most academically influential author in the field was Barbieri Carlo from the University of Milan, Italy, who had the highest number of publications (NP=5) and the highest H-index (H-index=5). Erythropoietin (ESA) is an effective clinical treatment option for anemia, a common complication of hemodialysis (HD) in CKD patients ^[3]^. In 2014, Barbieri Carlo’s team used AI-based reinforcement learning (RL) to optimize the ESA regimen, applying fit Q iteration (FQI) to compare it with the standard protocol, and the study revealed that the proportion of patients in the FQI group with hemoglobin in the target range increased by 27.6%, and the amount of medication required decreased by 5.13%. Although not experimentally validated, RL still has the potential to optimize or even replace the standard protocol ^[4]^. Another study by Barbieri Carlo developed an AI decision support system based on the anemia control model (ACM) for anemia care in HD patients. When the ACM recommendations were implemented, patients’ hemoglobin fluctuations significantly decreased, and compliance rates significantly increased, effectively reducing the use of ESAs and indirectly reducing the cost of treatment ^[5]^. In addition, Canaud Bernard from Franch and Cheung and Carol Y. from Hong Kong, China, have also contributed to the field (**Table 1**). Canaud Bernard proposed a strategy for automating digital health technologies (DHTs) in HD through DHTs. that drives automated HD through DHTs to remotely monitor patients, make medical decisions on real-time data, and provide health education as well as medical prescriptions ^[6]^. Cheung, Carol Y. developed a deep learning algorithm (DLA) based on AI to detect CKD through retinal images. The Singapore Epidemiology of Eye Diseases Study (SEED) was used for the development and validation of the DLA, and two datasets, the Singapore Prospective Study Program (SP2) and the Beijing Eye Study (BES), were used for external validation to assess the model performance in terms of the area under the receiver operating characteristic curve (AUC). The results showed that the AUC of the DLA model for retinal images was 0.911, which was significantly different from that of the SP2 and BES validation sets, suggesting that the DLA of retinal images can be used as an adjunctive or screening tool for CKD diagnosis ^[7]^.

National and institutional analysis showed that the USA and China have significant academic influence within the AI-CKD field, with NP and H-indexes greater than the mean of the top five corresponding data points (**Table 3**). The US authors Loftus and Tyler J. suggested that AI decision support systems can play a very important role within the field of nephrology by assisting in enhancing physicians’ decision-making abilities through objective prediction of results ^[8]^. Decision analysis based on AI can accurately predict the time frame of renal failure in CKD patients and risk factors for disease progression and can aid in the early diagnosis of the disease ^[9,10]^. A study by the Chinese author Liu Wei revealed that the KidneyOnline mobile application smart program was effective at improving renal outcomes in CKD patients by integrating and analyzing patient data and developing personalized care plans based on AI deep learning optical character recognition (OCR). The results showed that the KidneyOnline system significantly reduced patients’ mean arterial pressure and the risk of adverse renal outcomes compared to traditional care ^[11]^. Among the top five, the Mayo Clinic institution in the United States had the greatest number of publications (NP=7), and China had the most publications, followed by Tri-Service General Hospital, China Medical University, and The Chinese University of Hong Kong. The German institution, Fresenius Medical Care, had the greatest academic impact in the field (H-index=6). The authors at this institution, Bellocchio Francesco, developed the Prognostic Reasoning System for Chronic Kidney Disease (PROGRES-CKD) based on the naive Bayes classifier for predicting the outcome of adults with stage 3-5 CKD patients with end-stage kidney disease (ESKD) over a 6- to 24-month period. The study included 17,775 CKD patients at our institution and was externally validated using data from two independent cohorts. The results showed that PROGRES-CKD had a significant level of prediction at 6 months compared to the other two groups, while there was no significant difference in prediction level at 24 months, suggesting that PROGRES-CKD can be effective in clinical practice to assist physicians in making prognostic reasoning ^[12]^.

The results of keyword analysis revealed that artificial intelligence, chronic kidney disease, machine learning, prediction model, risk, deep learning and other keywords were popular (Figure 4b). Since the first study on AI within the field of CKD was published in 2005, there has been a gradual increase in AI research within the field centered on computer algorithms such as machine learning (ML), deep learning (DL), and predictive modeling. Two research outbreaks have occurred since 2018: on the one hand, computers objectify patient information data, avoiding the subjectivity of the human brain; on the other hand, computers have a more systematic and simpler way of processing a large number of data samples, which makes it easier for doctors to make clinical decisions based on objectified information data. Machine learning algorithms are able to process and analyze large amounts of medical information data and play an important role in disease diagnosis and prediction ^[13–15]^, personalized medicine and precision treatment ^[16]^, surgical planning and decision support ^[17]^, and electronic health record (EHR) analysis ^[18]^. In recent years, a variety of machine learning-based prediction models have been proposed for CKD diagnosis and risk prediction. Jiongming Qin established six machine learning models, including logistic regression and random forest models, based on the CKD dataset from the University of California, Irvine (UCI), to diagnose patients with CKD, and it was found that the diagnostic accuracy of the random forest model was 99.75%, which is more accurate than that of other models, and proposed the use of perceptron models to combine logistic regression and random forest models with a new comprehensive model, with an average diagnostic accuracy of 99.83% ^[19]^. In addition to random forests, models such as support vector machines (SVMs) and logistic regression have also shown good performance in the prediction of CKD. The combined use of SVM and logistic regression in one study resulted in a diagnostic accuracy of 99.75%. Another study proposed a noninvasive convolutional neural network-support vector machine (CNN-SVM) hybrid deep learning model for detecting urea concentrations in saliva for diagnosis, with a prediction accuracy of 96.59% ^[20]^. In addition, the machine learning model is also able to achieve noninvasive risk assessment of CKD and extract the information needed by physicians from complex clinical data for assisted diagnosis ^[18,21]^.

Cluster analysis of the keywords based on the timeline resulted in eight sets of cluster clusters, including retinal microvascular abnormality, renal failure, Bayesian network, anemia, MACHINE LEARNING and DEEP LEARNING, bone diseases, and allograft nephropathology. The prevalence of retinal microvascular abnormalities was greater in CKD patients, and significant changes in optical coherence tomography angiography (OCT-A) parameters of the retinal microvasculature were observed as the disease worsened ^[22,23]^. Varun Gulshan used 128,175 retinal images to construct deep CNN models for the automatic detection of retinopathy and macular edema and applied two independent external datasets for validation, which showed that deep CNN models based on deep machine learning algorithms are highly sensitive and specific for detecting retinopathy ^[14]^. In addition, semisupervised deep learning methods have also been applied in the classification and learning of medical images, providing some help in the auxiliary diagnosis of diseases ^[24]^. In recent years, ML has played an increasingly important role in the prediction and management of CKD. Compared with the traditional kidney failure risk equation (KFRE) model, the XGBoost model had an accuracy of 93.9% and specificity of 97.7% in predicting ESRD, which were significantly greater than those of the traditional model ^[25]^. The rotating forest (RotF) model also performed well in predicting CKD, with an accuracy and F-measure of 99.2% and an AUC value of 1.00 ^[26]^. Vinothini Arumugham constructed a prediction model for the early diagnosis of CKD using a deep neural network (DNN) with an adaptive moment estimation optimization function, and the results showed that the diagnostic accuracy of the DNN-CKD model was 98.75% ^[27]^. Bayesian networks (BNs) are a type of model that represents the probabilistic dependence between variables as a graphical model and can be analyzed by constructing multifactorial network relationships related to CKD and risk prediction ^[28]^. Xinfang Xie constructed a network model of the effects of ACEIs and ARBs on renal function as the primary outcome in CKD patients based on BNs to explore the preferred advantages of ACEI and ARB medications in CKD, and the results showed that ACEIs significantly reduced all-cause mortality in patients with CKD and that both drugs reduced the risk of renal failure ^[29]^. In addition, the relationships between various drugs or variables and CKD can all be meta-analyzed by constructing BN models ^[30–32]^. For patients suffering from CKD, anemia is one of the most common complications, and different algorithmic models based on AI, such as random forests and SVMs, are useful for the automated prediction of anemia to varying degrees ^[33]^. In the management of CKD-mineral and bone disorder (CKD-MBD) disease, the use of quantitative systems pharmacology (QSP) models combined with reinforcement learning (RL) has been shown to be fast and effective in achieving accurate treatment ^[34]^. Similarly, ML models have been able to predict the occurrence of possible outcomes after renal transplantation and to identify sets of genes associated with graft tolerance to guide immunotherapy by analyzing results from large amounts of data ^[35,36]^.

## 5. Conclusion

The continuous development of AI has led to the gradual adoption of various algorithmic models, including ML, DL, and CNN, in the medical field. In the field of chronic kidney disease (CKD), different algorithms play different roles in various fields, including early diagnosis of diseases, prediction of disease progression, prediction of related risk factors, and evaluation of drug safety and efficacy. The fusion algorithms and models of related research optimize existing prediction and evaluation schemes in this field to a certain extent, providing significant assistance for early diagnosis of the disease and standardized treatment. This study employed the bibliometric software VOSviewer and CiteSpace to visualize and analyze the relevant literature on the application of AI in the field of CKD. The objective is to identify research hotspots and cutting-edge development trends in this field, thereby providing a foundation for future research.

## Data Availability

All relevant data are within the manuscript and its Supporting Information files.

## Acknowledgments

This work was supported by grants from the National Natural Science Foundation of China (grant no. 81873343), the National Natural Science Foundation of China Youth Fund (grant no. 82205190).

